# Rare coding variants in schizophrenia-associated genes affect generalised cognition in the UK Biobank

**DOI:** 10.1101/2023.08.14.23294074

**Authors:** Eilidh Fenner, Peter Holmans, Michael C O’Donovan, Michael J Owen, James T R Walters, Elliott Rees

## Abstract

Cognitive impairments in schizophrenia are associated with poor outcomes and are largely unimproved by current medications. It remains uncertain to what extent cognitive impairments arise from shared aetiology and biology with schizophrenia, or are a consequence of having the condition. We analysed exome-sequencing data from 76,783 UK Biobank participants without schizophrenia to test for association between generalised cognition (*g*) and rare (minor allele count≤5) variants in schizophrenia-associated genes. Protein-truncating and deleterious missense variants in loss-of-function intolerant genes were associated with lower *g*. Stronger effects on *g* were found for protein-truncating variants in genes implicated in schizophrenia by rare coding variation, and for deleterious missense variants in credible causal genes at schizophrenia common allele loci. These findings indicate that biological processes disrupted in schizophrenia by common and rare variants are associated with *g* in unaffected individuals, suggesting the relationship between impaired cognition and schizophrenia reflects, in part, a shared underlying biology.

**Graphical Abstract:** Figure 1
Graphical abstract. RCVs = rare coding variants. QC = quality control. *g =* generalised cognition.

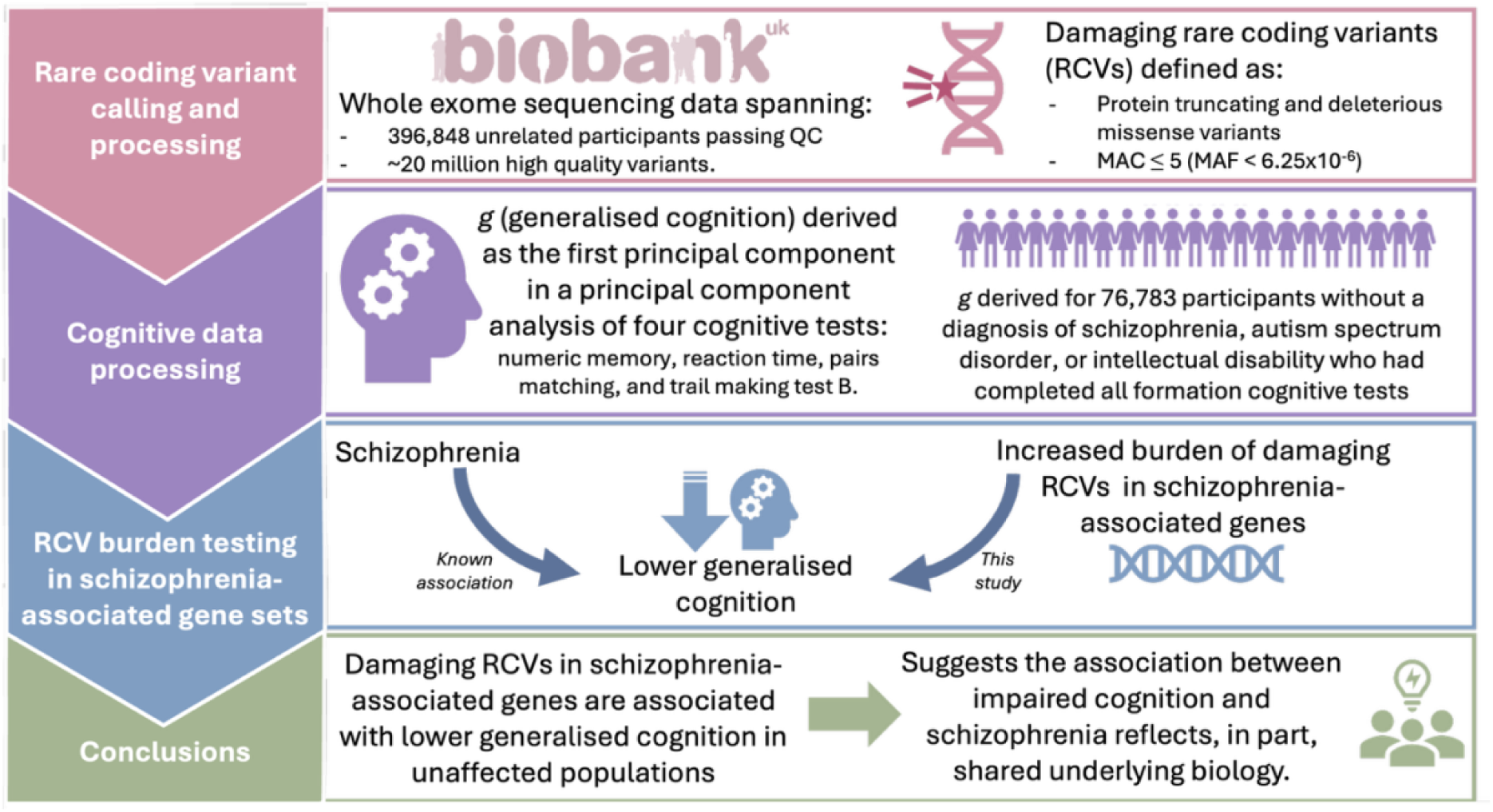

## Introduction

Schizophrenia is a severe and clinically heterogeneous psychiatric disorder with a lifetime prevalence of just under 1%^1,2^. Impaired cognitive function is an important feature of schizophrenia that often precedes the onset of psychosis^3–5^ and is a strong predictor of functional outcomes, encompassing social, occupational, educational, and interpersonal functioning, the ability to live independently, and the ability to engage in day-to-day activities^6,7^. Relative to controls, those with schizophrenia demonstrate impairments across multiple cognitive domains, suggesting the disorder is associated with a generalised, rather than a domain specific, cognitive impairment. Indeed, measures of generalised cognition, or *g*, in individuals with schizophrenia are on average around 1.5 standard deviations lower than in controls^8–10^. Furthermore, *g* explains the majority of variance in cognition between people with schizophrenia and healthy controls, and is also a better predictor of functional outcome in schizophrenia than any individual cognitive test^11,12^.

Schizophrenia is highly heritable and polygenic, with liability conferred by rare and common alleles in many genes, particularly those under selective constraint against mutations predicted to result in a loss of protein function, also known as loss-of-function intolerant (LoFi) genes^13–15^. The largest genome-wide association study (GWAS) of common alleles (minor allele frequency (MAF) > 1%) in schizophrenia to date identified 287 distinct associated loci^16^, and among these, fine-mapping prioritised 106 protein-coding genes as credibly causal. The genome-wide burden of rare copy number variants (CNVs) is also increased in schizophrenia cases compared with controls, with a set of 63 CNVs reported to be associated with developmental disorders showing particular enrichment in cases^17,18^. Moreover, 13 of these CNVs have been robustly associated with schizophrenia liability^17,18^. Finally, sequencing studies have identified 12 genes that are individually enriched in schizophrenia for damaging types of rare coding variants (RCVs)^19,20^. There is also evidence for a convergence between genes implicated in schizophrenia by common and rare alleles^16,19^.

Genetic liability for schizophrenia is pleiotropic, with shared effects on liability to several other psychiatric and developmental disorders as well as on variation in cognitive function in the general population. For example, the 13 CNVs robustly associated with schizophrenia are all risk factors for developmental disorders^17^, and individuals without a psychiatric or developmental disorder who carry one of these CNVs perform worse on a range of cognitive tests compared to those who do not have such CNVs^21,22^. Similarly, damaging RCVs in developmental disorder-associated genes are associated with poorer cognition in individuals without a psychiatric or developmental disorder^23,24^. Finally, a higher burden of schizophrenia common alleles is associated with lower cognitive function in the population^25,26^.

Less is known about the effects of damaging RCVs in schizophrenia genes and loci on cognition in individuals without a diagnosis of schizophrenia, autism spectrum disorder (ASD) or intellectual disability (ID). Studying these individuals helps isolate the direct effects of rare variant liability for schizophrenia on cognition, independent of the effects of other factors related to the pathophysiology of the disorder, or that emerge as a consequence of schizophrenia or other neurodevelopmental disorders (NDDs) (e.g. antipsychotic medication use, long-term symptoms, or lifestyle factors). This reduces the risk of reverse causation, whereby differences in cognitive ability may be a consequence of the disorder and/or its treatment. It is therefore important to establish whether damaging RCVs in schizophrenia-associated genes impact cognition in those unaffected by this disorder, as this would provide evidence that the association of impaired cognition with schizophrenia is, at least in part, explained by shared biology between schizophrenia and cognitive function.

Recent studies using data from the UK Biobank (UKBB) have shown damaging RCVs in LoFi genes are associated with longer reaction-time, lower fluid intelligence scores, and lower levels of educational attainment^23,27^. One study also reported that damaging RCVs in genes mapping to schizophrenia loci identified by GWAS are significantly associated with lower educational attainment, and also nominally significantly associated with longer reaction-time and lower fluid intelligence^23^. However, that study did not determine whether this association was attributable to those loci being associated with schizophrenia, or whether it was confounded by the fact that LoFi and brain-expressed genes are enriched both within these schizophrenia loci^15^. It also did not test the specific genes within those loci that have been reported to be credibly causal. There has also been no study examining the relationship between cognition in individuals without a psychiatric or developmental disorder and RCVs in sets of genes implicated in schizophrenia by rare CNVs or RCVs.

In the current study, we analysed exome sequencing data from 396,848 unrelated participants passing quality control (QC) in the UKBB to examine whether damaging RCVs in genes implicated in schizophrenia have effects on cognition in people without a diagnosis of schizophrenia, ASD, or ID (full study design presented in Methods). Whereas previous RCV studies of cognition in the UKBB analysed only individual tests of cognition^23,24^, we analysed a measure of generalised cognition (*g*), which we formed from a principal component analysis of four cognitive tests. We focussed our study on *g*, instead of individual cognitive tests, as it is a more robust measure of general cognitive ability, and also because schizophrenia is strongly associated with a generalised rather than a specific cognitive deficit^12,28,29^. Our findings demonstrate that rare protein-truncating variants (PTVs) and damaging missense variants in LoFi genes are associated with lower *g*, which to our knowledge has not been shown before. We also show that rare PTVs in genes implicated in schizophrenia by RCVs have a significantly stronger effect on *g* than that observed for rare PTVs in LoFi genes and brain-expressed genes. Additionally, rare deleterious missense variants in credible causal genes underlying schizophrenia common allele loci were associated with lower *g,* and this effect was significantly stronger than that observed for rare deleterious missense variants in all genes mapping to the schizophrenia common allele loci, and for rare deleterious missense variants in LoFi genes or brain-expressed genes.

Overall, these findings indicate that biological processes disrupted in schizophrenia by common and rare variants are linked to generalised cognitive function in unaffected individuals. This suggests the association of impaired cognition with schizophrenia reflects shared underlying biology between schizophrenia and cognitive function.

## Results

### Distribution of protein-coding variation

19,724,617 high-quality autosomal protein coding variants were observed in 396,848 unrelated UKBB participants that passed quality control (QC). We defined rare variants as those with a minor allele count (MAC) ≤ 5 (equating to a MAF < 6.25 × 10^−6^), a definition similar to that of other UKBB rare variant studies^23,24^. 75.6% of observed variants were rare. Substantial variation was observed in the number of rare variants per participant across genetic ancestries (Supplementary Table 1; see Supplementary Materials 1.1 for detail on ancestry inference), with participants that were genetically similar to the 1000 Genomes^30^ European superpopulation (1KGP-EUR-like) carrying fewer rare damaging variants (mean number of rare PTVs carried = 2.02) compared with participants genetically similar to other 1KGP superpopulations (mean number of rare PTVs carried ranging from 5.80 to 8.51; Supplementary Table 1). This difference was expected given allele frequencies were defined by the UKBB sample itself, and 95% of the cohort were of 1KGP-EUR-like genetic ancestry. We controlled for differences in the rates of RCVs across populations by analysing participants in each genetically inferred ancestry group separately.

### Estimating generalised cognition in the UKBB

Generalised cognition (*g*) was estimated in 76,783 participants that passed QC (see Methods and Supplementary Materials 1.2 for a full description of how *g* was derived). In these participants, *g* explained 38.9% of the variance of the tests included in its formation (numeric memory, reaction time, pairs matching, and trail making test B; see Supplementary Table 2 for a description of these tests). As expected*, g* was moderately correlated with each of the individual tests used in its derivation (weakest correlation coefficient = −0.53, Supplementary Table 3), as well as with cognitive tests not used to derive *g* (weakest correlation co-efficient = 0.41, Supplementary Table 3). All correlations of *g* and individual cognitive tests were in the expected direction, where higher *g* correlated with higher score on each test (we note that negative correlation coefficients are expected when lower scores for a given test indicate higher cognitive ability). These results suggest *g* forms a cross-domain measure of generalised cognition. We performed a sensitivity analysis of *g* by modifying some of the individual cognitive tests included in its formation (see Supplementary Materials 1.3 for details) and found alternative estimates of *g* were highly correlated with the original estimate (weakest correlation co-efficient = 0.8, Supplementary Table 4), suggesting *g* is robust to different input measures.

### Rare coding variants in schizophrenia-associated genes and generalised cognition

We investigated the relationship between damaging RCVs in schizophrenia-associated genes and *g* in UKBB participants without a diagnosis of schizophrenia, ASD, or ID (see Supplementary Figure 1 for a summary of analyses). We focussed our analysis on the following classes of rare variant: PTVs (splice acceptor, splice donor, stop-gain and frameshift variants predicted with high-confidence to cause loss of protein function^31,32^); deleterious missense variants (REVEL^33^ scores > 0.75); and synonymous variants. We investigated the impact on cognition of these variants in LoFi genes defined by GnomAD pLI scores^34^ ≥ 0.9, and in sets of genes implicated in schizophrenia by the largest RCV^19^, common allele GWAS^16^, and CNV studies^17,18^ of the disorder (see Methods and Supplementary Table 5 for a description of the gene sets tested).

#### Constrained and brain expressed genes

In UKBB participants that were genetically similar to the 1KGP^30^ European superpopulation, rare PTVs and deleterious missense variants in LoFi genes were associated with lower *g* (rare PTVs: β = −0.062, p = 3.3 × 10^−17^; rare deleterious missense variants: β = −0.040, p = 6.3 × 10^-9^; Figure 2; Supplementary Table 6). Rare synonymous variants in LoFi genes were not associated with *g* (Figure 2; Supplementary Table 6). 17.3% of the sample carried at least one rare PTV in a LoFi gene, and 20.5% carried at least one rare deleterious missense variant in a LoFi gene (Supplementary Table 7). A greater number of rare PTVs and deleterious missense variants carried in LoFi genes was associated with lower *g* (Supplementary Figure 2). Similar effects on *g* were observed for RCVs in LoFi genes in males and females (Supplementary Figure 3). For all other genetic ancestry groups tested, no significant effects on *g* were observed for any class of RCV in LoFi genes (Supplementary Table 8), most likely due to small sample sizes. Therefore, all analyses presented hereafter are based on participants that are genetically similar to the 1KGP^30^ European superpopulation.

**Figure 2.**
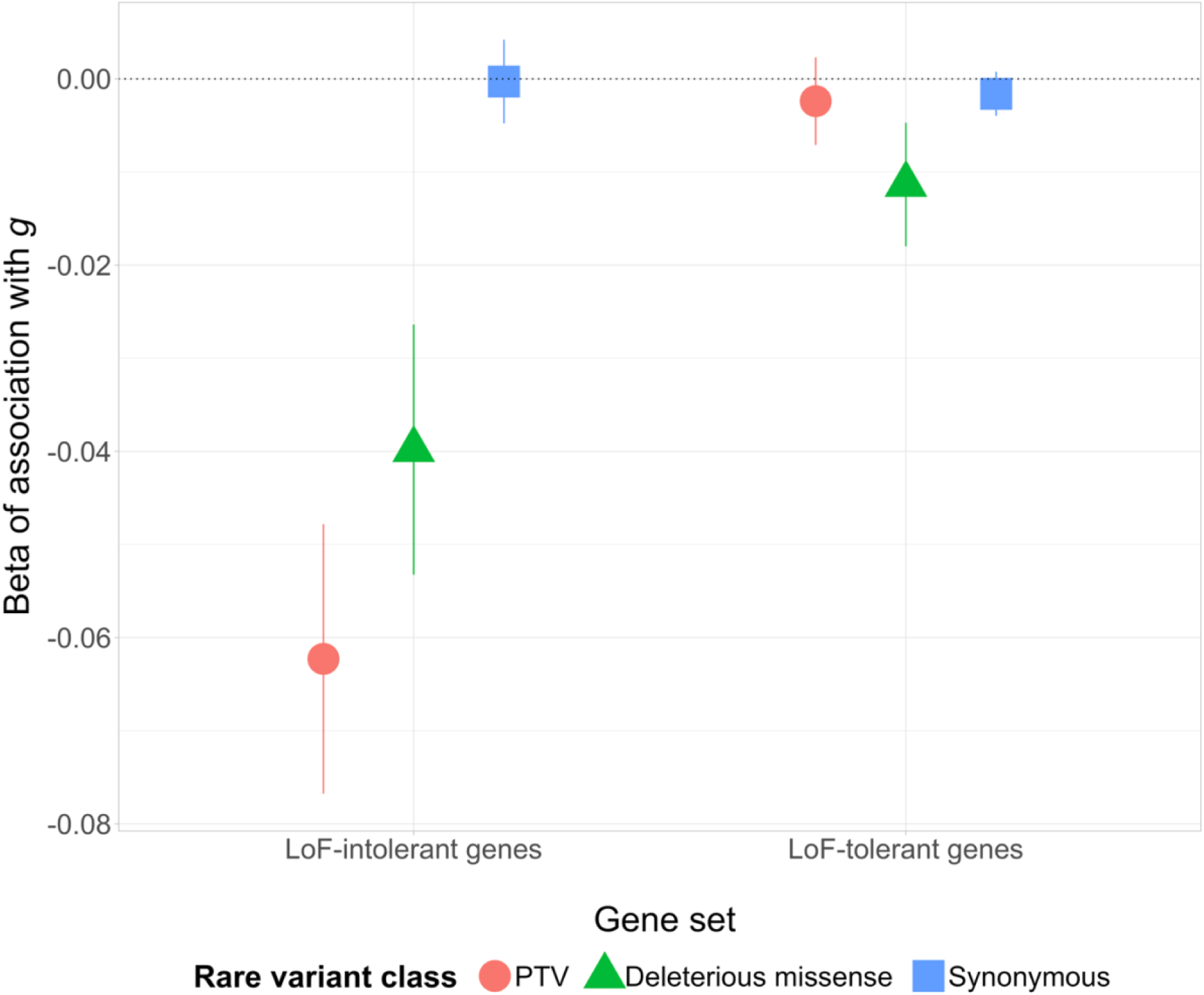
Association between *g* and different classes of rare coding variants in LoF-intolerant and LoF-tolerant genes. Error bars display 95% confidence intervals. LoF = loss-of-function; PTV = protein-truncating variant; *g* = generalised cognition.

The associations of *g* with rare PTVs and rare deleterious missense variants in LoFi genes remained significant when both classes of RCVs were modelled together and after including schizophrenia-enriched CNVs and schizophrenia polygenic risk scores (PRS) in the model (Supplementary Table 9). This suggests that the effect on cognition of each of these classes of genetic variation are largely independent from one another.

Rare PTVs and rare deleterious missense variants in brain expressed genes (as defined in Supplementary Table 5) were associated with lower *g* (PTVs: β = −0.018, p = 3.3 × 10^-8^; deleterious missense variants: β = −0.020, p = 1.1 × 10^-6^; Supplementary Table 6). Rare PTVs in LoFi genes that were brain expressed had significantly stronger effects on *g* than LoFi genes that were not (z-test p = 0.0099). There was no significant difference in the effects on *g* between rare deleterious missense variants in brain expressed and non-brain expressed LoFi genes (z-test p = 0.47).

Rare PTVs and synonymous variants in LoF-tolerant genes were not associated with *g* (Figure 2; Supplementary Table 6). Rare deleterious missense variants in LoF-tolerant genes were associated with lower *g* (β = −0.011, p = 0.0008; Figure 2; Supplementary Table 6), although the effect size for these variants was significantly weaker than that observed for deleterious missense variants in LoFi genes (z-test p value = 9.8 × 10^-5^).

To assess whether RCVs in LoFi genes have effects on generalised cognitive ability beyond those captured by individual cognitive tests, we included individual cognitive test scores as covariates in the analysis of *g* with RCV burden in LoFi genes. Controlling for individual cognitive tests that were not included in the formation of *g* (trail-making test A, fluid intelligence, and symbol digit substitution), as well as those that were (numeric memory, reaction time, pairs matching, and trail making test B; see Supplementary Table 2 for a description of all tests), only partly attenuated the significant association between rare PTVs and deleterious missense variants in LoFi genes and *g* (Supplementary Table 10). These findings suggest that damaging RCVs in LoFi genes have broad effects on cognition that extend beyond those captured by any individual cognitive test.

#### Genes associated with rare coding variants in schizophrenia

We defined genes associated with RCVs in schizophrenia as those implicated in the disorder at a false discovery rate (FDR) < 5% in the Schizophrenia Exome Sequencing Meta-Analysis Consortium (SCHEMA) study^19^ (n autosomal genes = 29), hereafter referred to as SCHEMA FDR < 5% genes. Rare PTVs in this set were associated with lower *g* (β = −0.24, p = 0.0004) but rare deleterious missense were not (Figure 3, Supplementary Table 11). PTVs in SCHEMA FDR < 5% genes were associated with *g* with a similar beta (β = −0.24) to that of schizophrenia-enriched CNVs (β = −0.28, Supplementary Table 9). The effect on *g* for rare PTVs in SCHEMA FDR < 5% genes was significantly greater than that observed for rare PTVs in all LoFi genes and brain expressed genes (respective z-test p-values = 0.0038 and 0.00045).

**Figure 3.**
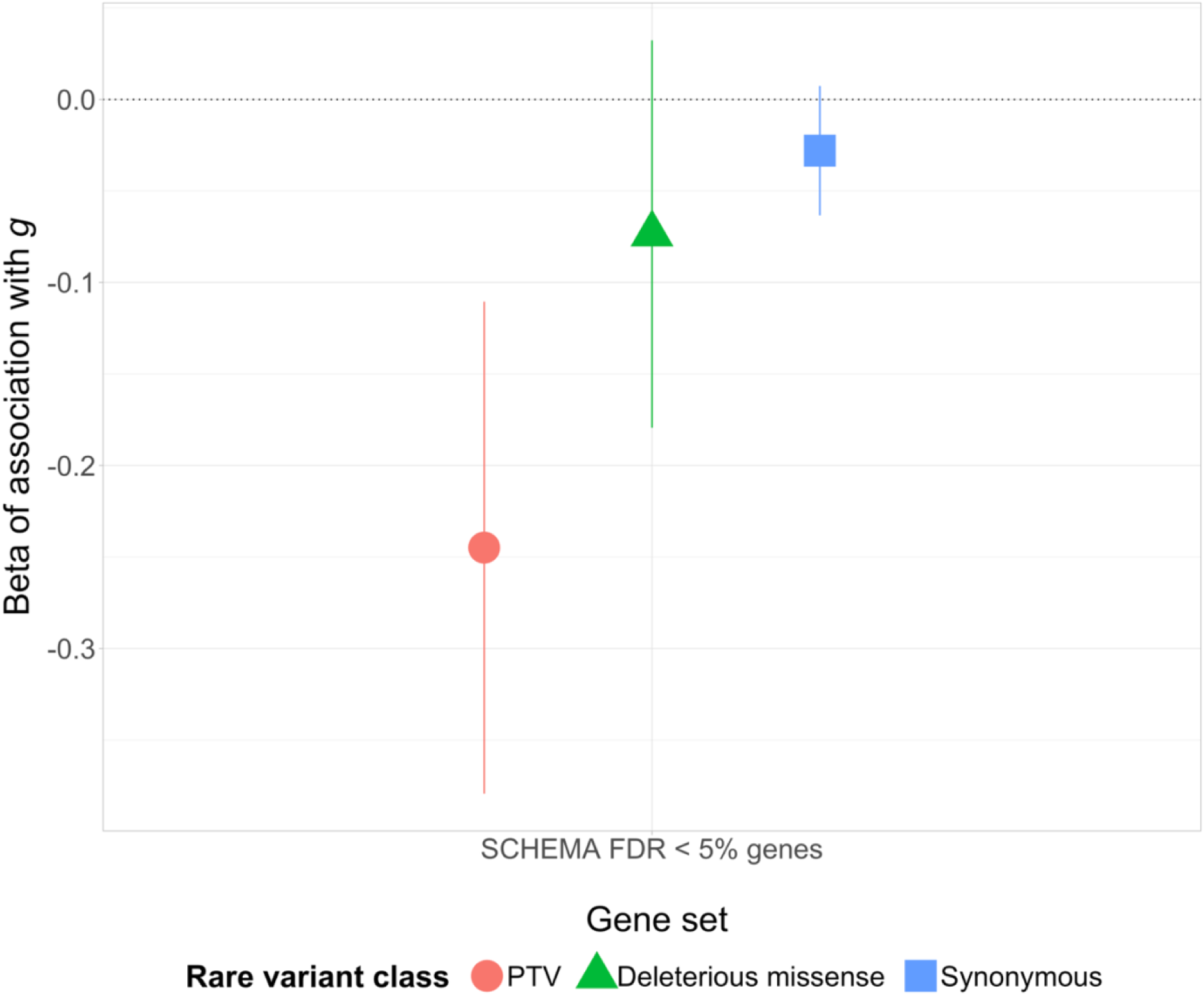
Association between *g* and different classes of rare variants in SCHEMA FDR < 5% genes from ^19^. Error bars display 95% confidence intervals. FDR = false discovery rate; PTV = protein-truncating variant; g = generalised cognition.

#### Schizophrenia common allele loci

The largest schizophrenia GWAS of common variation to date^16^ identified 282 associated autosomal loci and prioritised credible causal protein-coding genes for some of these loci. We analysed RCVs in the following schizophrenia common allele loci gene sets: 1) a broad set including all genes overlapping the common variant loci (n = 1,715); 2) genes closest to the index SNP for each locus (n=186); 3) and genes prioritised as credible causal in that paper^16^ (n = 101).

Neither rare PTVs nor rare deleterious missense variants in genes mapping to broad schizophrenia common variant loci were associated with *g* (Figure 4, Supplementary Table 12). At genes nearest the locus index SNP, rare deleterious missense variants (β = −0.073, p = 0.0045) but not rare PTVs were associated with *g* (Supplementary Table 12). The same was true for the credible causal genes (rare deleterious missense variants β = −0.12, p = 0.0005; Figure 4; Supplementary Table 12). The effect on *g* of rare deleterious missense variants in credible causal genes was significantly greater than in non-prioritised genes at GWAS loci (z-test p = 0.0008). It was also greater than the effect of rare deleterious missense variants in LoFi genes in general (z-test p = 0.010), which is important to demonstrate since schizophrenia common alleles are enriched for LoFi genes^15^.

**Figure 4.**
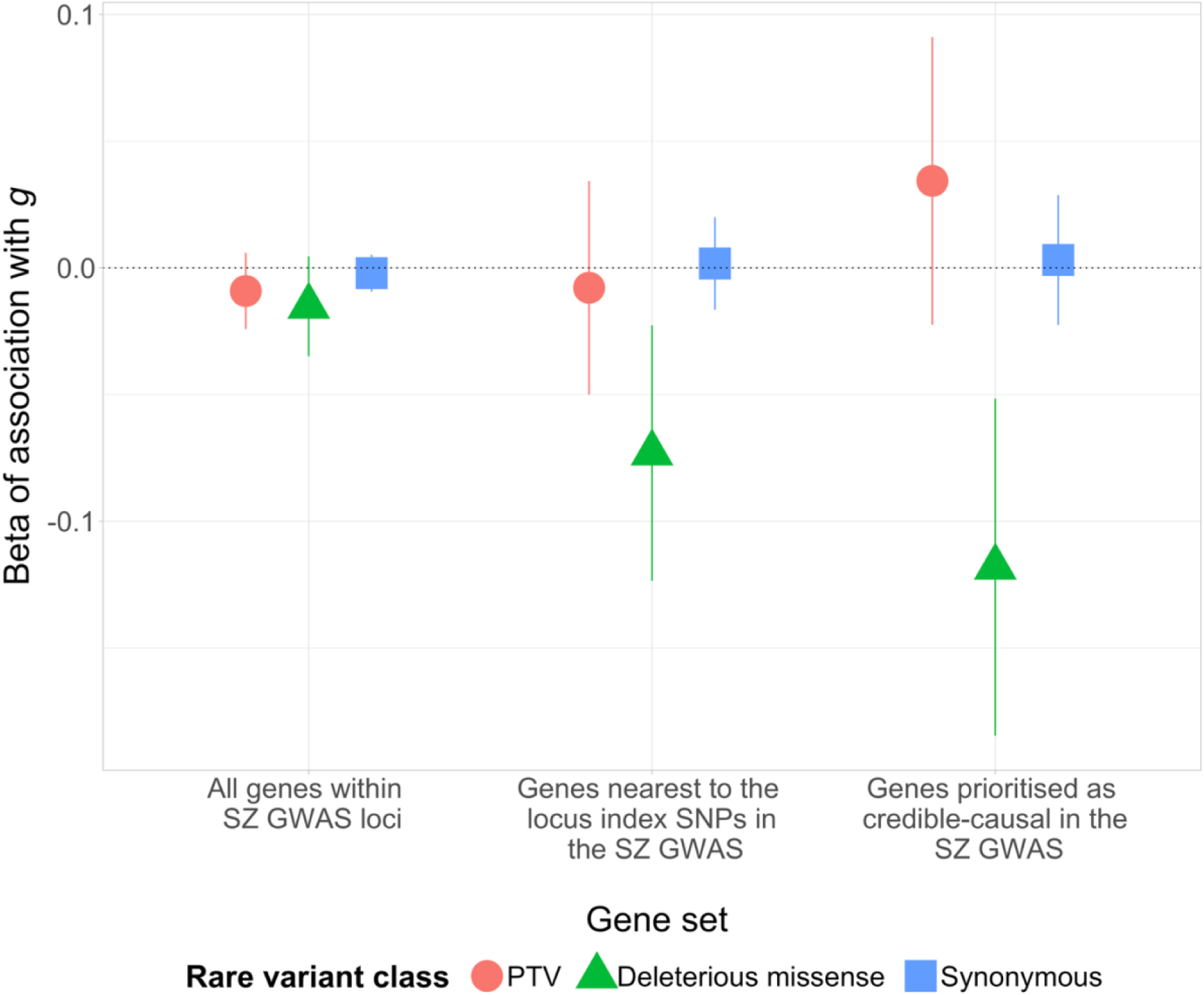
Association between *g* and different classes of rare variants in various schizophrenia (SZ) common allele loci/ GWAS sets, from ^16^. Error bars display 95% confidence intervals. PTV = protein-truncating variant; g = generalised cognition.

Within the credible causal set, rare deleterious missense variants in the 61 fine-map prioritised genes (β = −0.12, p = 0.0029) and in the 44 SMR prioritised genes (4 of which were also prioritised by fine-mapping, β = −0.12, p = 0.046) had similar effect sizes (Supplementary Figure 4; Supplementary Table 12). Prioritisation as a credible causal candidate by SMR^16^ required a gene to have an brain eQTL and therefore brain expression could act as a confounder. The effects on *g* for rare deleterious missense variants in credible causal genes, and also in the subset of SMR prioritised genes, were significantly stronger than in all brain-expressed genes that were not part of these respective sets (credible causal genes vs. brain-expressed genes: z-test p = 0.0018; SMR prioritised genes vs. brain-expressed genes: z-test p = 0.048). Thus, the associations we observe here do not simply reflect a background association between *g* and rare deleterious missense variants in brain-expressed genes.

#### Schizophrenia CNV loci

RCVs in genes mapping to the following sets of CNVs implicated in schizophrenia were tested for association with *g*: 1) schizophrenia-enriched CNVs (n genes = 886), which includes 63 developmental disorder-associated CNVs that are collectively enriched in schizophrenia^17^; 2) schizophrenia-associated CNVs (n genes = 178), which includes 13 CNVs with strong evidence for association with schizophrenia. See Methods and Supplementary Table 5 for further details on these CNV gene sets.

No class of RCV in the 886 genes mapping to schizophrenia-enriched CNVs was associated with *g* (Figure 5; Supplementary Table 13). Rare PTVs in LoFi genes within the schizophrenia-enriched CNV loci (n = 144) were associated with lower *g* (β = −0.086, p = 0.0099, Figure 5, Supplementary Table 13) but the effect size did not significantly differ from that in LoFi genes outside CNV loci (z-test p = 0.23). No class of RCV in LoF-tolerant genes within the schizophrenia-enriched CNV loci was associated with *g* (Figure 5, Supplementary Table 13).

**Figure 5.**
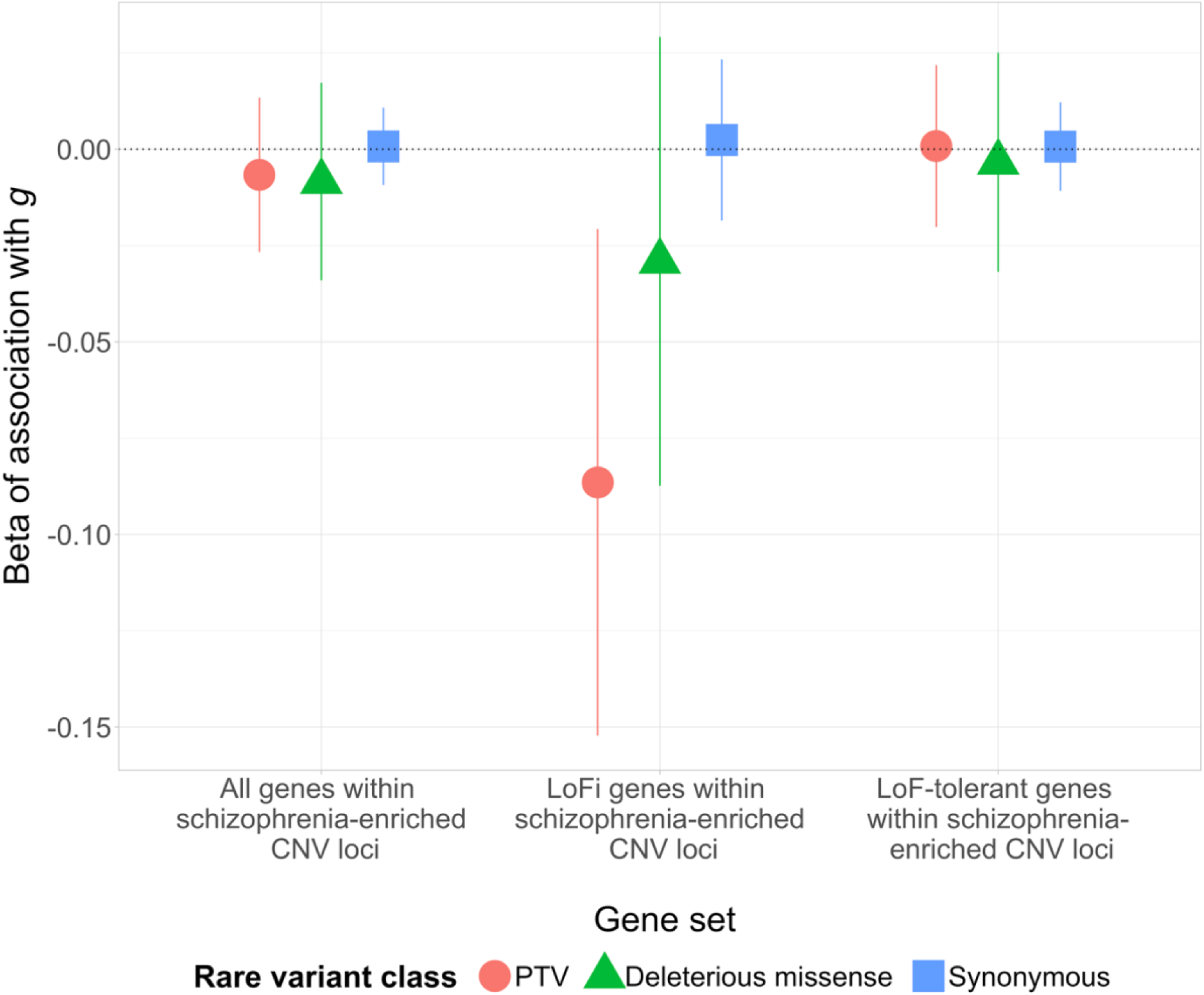
Association of *g* with different classes of rare variants in genes within schizophrenia-enriched CNV loci. LoFi and LoF-tolerant genes in schizophrenia-enriched CNV loci were also tested separately. Error bars display 95% confidence intervals. LoFi = LoF-intolerant; PTV = protein-truncating variant; *g* = generalised cognition.

No associations with *g* were observed for any class of RCV within the 178 genes mapping to the schizophrenia-associated CNV set or in the subsets of those genes that were LoFi or LoF-tolerant (Supplementary Table 13).

### Single gene burden tests

Single-gene association tests were performed for three classes of RCVs (PTVs, deleterious missense variants, and a combined (PTV + deleterious missense) variant class) in 1,003 unique genes taken from the SCHEMA FDR < 5% gene set, the credible causal schizophrenia GWAS gene set, and genes within schizophrenia-enriched CNV loci (number of single gene tests = 3,009). We did not test genes with fewer than five qualifying variants due to a lack of power. No gene showed significant evidence for association after Bonferroni correction (corrected p value 0.05/3,009 = 1.66 × 10^-5^; Supplementary Table 14).

## Discussion

In the UKBB, we show that damaging RCVs in genes and loci implicated in schizophrenia by common variant and RCV studies are associated with lower *g* in participants without a diagnosis of schizophrenia, ASD or ID. To our knowledge, this is the first study to investigate the relationship between RCVs in schizophrenia genes and *g*. We focussed on *g*, as opposed to individual cognitive tests, for two main reasons. First, a large body of evidence suggests schizophrenia is associated with global cognitive impairment^12,28,29^, and compared to controls, people with schizophrenia show greater impairment in *g* than for tests of specific cognitive domains^35^. Second, compared to individual cognitive tests, *g* reduces noise by limiting test-specific error and is a robust measure of general cognitive ability^36–38^.

Genes under selective constraint for PTVs have consistently been shown to be enriched for common alleles, rare CNVs, and RCVs in people with schizophrenia^13–15^. Here, we provide two novel lines of evidence demonstrating that damaging RCVs in these LoFi genes also contribute to variation in *g* in people without a diagnosis of schizophrenia, ASD, or ID. We first show that rare PTVs and deleterious missense variants in LoFi genes are significantly associated with lower *g,* and that these associations remained significant when adjusting for CNVs and common alleles that confer liability for schizophrenia and are known to influence cognition in unaffected individuals. This suggests the impact of RCVs on *g* is at least partially independent of these other genetic factors. We then show that the association of *g* with RCVs in LoFi genes is not explained by any individual cognitive test, including reaction time and fluid intelligence, both of which have recently been associated with damaging types of RCVs in LoFi genes^23,27^. These findings suggest the impact of these variants extends beyond any individual cognitive domain, and that they have a broad, generalised effect on cognitive function. Such a generalised effect of these RCVs suggests they influence brain mechanisms or systems that control processes central to cross-domain cognition.

RCVs in LoFi genes are associated with both schizophrenia and cognition, but there are many LoFi genes and whether the same LoFi genes have pleiotropic effects on schizophrenia and cognition in unaffected individuals is unknown. We show for the first time that carriers of rare PTVs in genes associated with RCVs in schizophrenia have significantly lower *g*, and that the effect of rare PTVs in SCHEMA FDR < 5% genes on *g* is significantly greater than that observed for rare PTVs in LoFi genes and in brain expressed genes. This finding is consistent with genic pleiotropy between schizophrenia and general cognitive function in unaffected individuals. We note that carriers of rare PTVs in SCHEMA FDR < 5% genes had a 0.24 standard deviation decrease in *g*, which is similar to the reduction in *g* observed for carriers of schizophrenia-enriched CNVs (0.28 standard deviations).

We also provide novel evidence that rare deleterious missense variants in credible causal genes within schizophrenia-associated common variant loci are associated with lower *g*, and that this is not confounded by the fact that this set of genes is enriched for LoFi genes and those that are brain expressed. These findings advance previous work that only examined the effects on cognitive traits of RCVs in broadly defined schizophrenia common allele loci, and did not control for the effects of constraint or brain expression^23^. In addition, our analysis of schizophrenia credible causal genes also provides orthogonal support for the SMR and fine-mapping approaches used to prioritise credible causal genes for schizophrenia in the study by Trubetskoy et al.^16^, suggesting data from population studies of RCVs could assist gene prioritisation within GWAS loci.

By finding the strongest effects on *g* for RCVs in genes that have individually been implicated in schizophrenia (i.e. the credible causal genes and RCV associated gene sets), our study supports the hypothesis that the biology of schizophrenia overlaps that for general cognitive function in the population. Highlighting these specific gene sets in which deleterious mutations have particularly strong effects on cognition allows us to more definitively highlight a role of these schizophrenia-associated genes in cognitive impairment. Moreover, these sets of genes have plausible biological links to cognition, given they are enriched for genes encoding proteins related to the synapse^16,19^, which has important roles in learning and memory^39^.

CNVs associated with developmental disorders have previously been shown to be collectively enriched in schizophrenia^17^ and associated with lower scores on individual cognitive tests in unaffected individuals^21,22^. In the current study, we mirror findings from other cohorts demonstrating lower generalised cognition in carriers of these CNVs^40^. We also found that rare PTVs in LoFi genes overlapping the CNV loci are associated with lower *g*, however, these effects were not significantly different from those observed for rare PTVs in all LoFi genes, contrasting with findings based on other schizophrenia gene sets. We speculate that this reflects the highly polygenic nature of schizophrenia^41^, and the multigenic nature of all but one schizophrenia-associated CNV, a consequence of which would be that unless each CNV contains a large number of schizophrenia-causal genes, the proportion of such genes in CNVs is unlikely to be substantially elevated over LoFi genes in general.

The favourable psychometric properties of *g* compared to individual cognitive tests^37,38^ is a strength of our study, but the associated requirement for multiple cognitive test scores will almost certainly induce a participation bias that would lead to under-representation of mutations that reduce the likelihood of completing all cognitive tests. This is plausible since previous work has shown that UKBB participants with PTVs in genes associated with ASD or in LoFi genes are less likely to have completed cognitive testing^42^. The known UKBB volunteer bias, whereby participants have a higher average socio-economic status and are generally healthier than the UK population^43^ is likely to have a similar effect. Both biases would result in underestimation of the effect sizes of variants that impact *g*, but are unlikely to change the general direction of observed effects^44^. Future research focused on understanding the relationship between different classes of RCVs and participation in the voluntary and cognitive components of the UKBB could inform the interpretation of findings from this and wider studies.

While our findings show that damaging RCVs in schizophrenia-associated genes are associated with lower *g* in the general population, the effect sizes we observe may differ in clinical populations, as we studied participants without a diagnosis of schizophrenia, ASD, or ID to reduce the likelihood of our findings reflecting reverse causation. Another limitation this study faced, despite the large sample size, was that some analyses were underpowered; for example, we lacked power to implicate individual genes in cognition. We were also underpowered to analyse RCVs in non-1KGP-EUR-like genetic ancestries. It is important for future work to examine the relationship between damaging RCVs in schizophrenia-associated genes and loci and cognitive function in clinical populations and in more diverse and representative samples.

In conclusion, we show for the first time that damaging RCVs in LoFi genes contribute to variation in generalised cognitive function in UKBB participants without a diagnosis of schizophrenia, ASD, or ID, with significantly stronger effects on cognition observed for damaging RCVs in genes previously associated with schizophrenia. These findings suggest that biology impacted in schizophrenia by common and rare alleles is associated with cognitive function in the population, and that the genetic overlap between schizophrenia and cognition in unaffected individuals is not explained by general gene properties such as LoF-intolerance or brain expression. They also suggest that the association of impaired cognition in schizophrenia is, at least in part, explained by shared biology between schizophrenia and cognitive function. Our study demonstrates the utility of exploiting large sequencing datasets of unaffected individuals, such as the UKBB, to identify genes with shared effects on cognition and schizophrenia and provides a route towards determining the biological processes underlying cognitive impairment in the disorder.

## Methods

### Key Resources Table

**Table 1.**
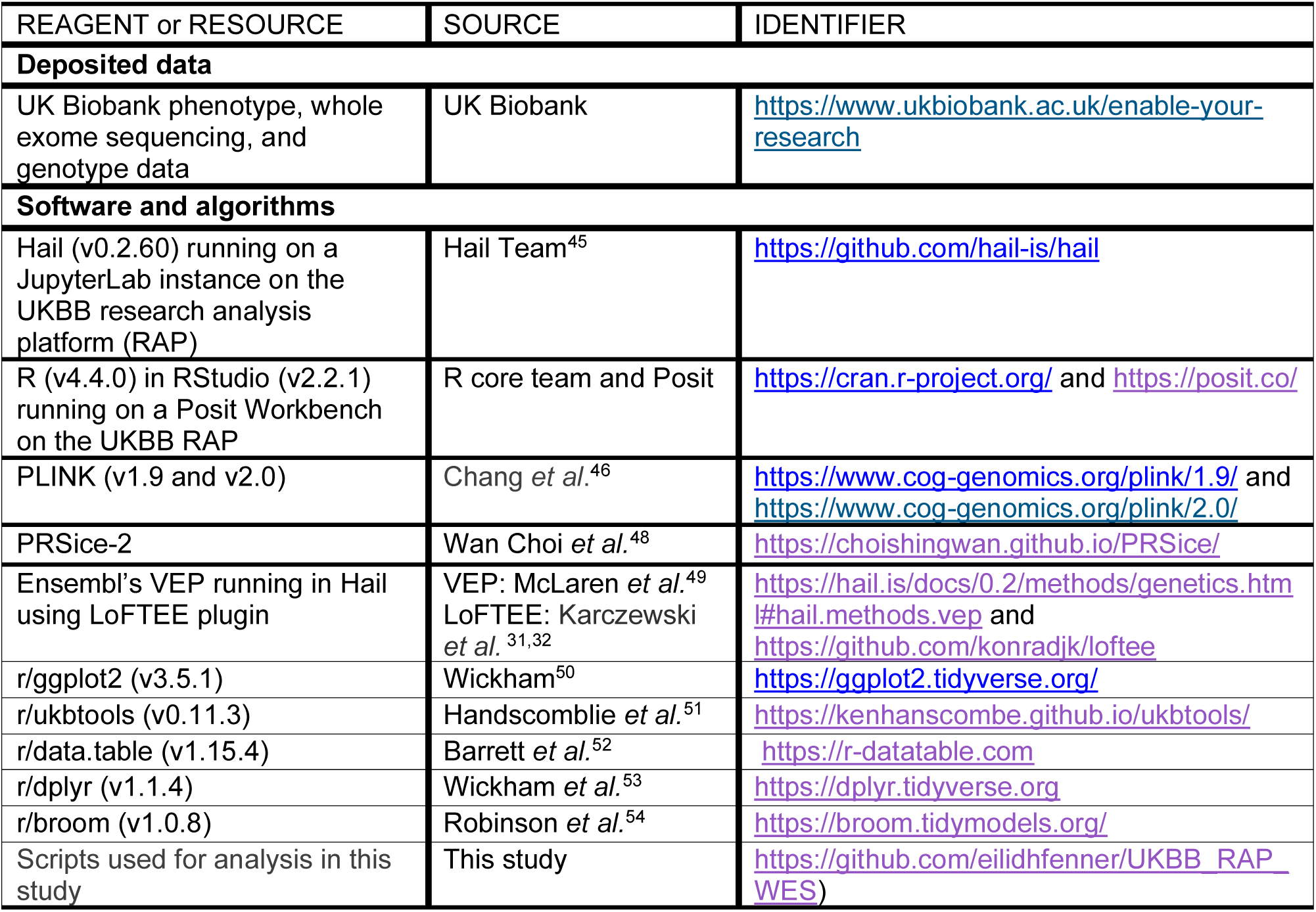
Key resources table.

### Method details

### UK Biobank

Between 2006 and 2010, the UKBB recruited around 500,000 participants from the UK through NHS registers, with no exclusion criteria apart from reasonable access to an assessment centre. Participants were aged 40-69 at recruitment and 54% were female. All participants gave consent for their data to be used by UKBB projects and agreed to being followed up (Supplementary Materials 1.4). The research presented in this study was conducted using the UKBB resource under Application Number 13310.

### Whole exome sequencing data

Whole exome sequencing data for 469,385 UKBB participants were generated using the IDT xGen Exome Research Panel v1.0 exome capture kit and Illumina NovaSeq 6000 instruments; S2 flow cells were used for the initial 50k sequencing release, and S4 flow cells used for all following samples. Further details on sequencing protocol can be found in the primary UKBB publication^55^.

#### Data processing and quality control

WES data was accessed and analysed using the UKBB Research Analysis Platform (RAP). Data was initially accessed as a joint-genotyped variant call set in pVCF format, which was centrally generated by the UKBB with Deep Variant 0.010^56^ using CRAMs that were processed using the ‘OQFE protocol’^57–60^, with reads aligned to GRCh38 reference genome^61^. For further details, see ^62,63^.

We performed genotype, sample, and variant QC using Hail (version 0.2.60)^45^ running on a JupyterLab instance on the RAP and R (using RStudio v2.2.1) running on a Posit Workbench on the RAP. The original dataset spanned 22,854,479 variants and 469,385 individuals. Multiple steps of variant- and genotype-level QC were undertaken to ensure only high-quality variants remained in the dataset. The number of variants excluded and retained following each QC step are presented in Supplementary Table 15. Genotypes were excluded if they met any of the following criteria: depth ≤ 10x; genotype quality ≤ 30; homozygous genotypes for the reference allele with an allele balance > 0.1; homozygous genotypes for the alternate allele with an allele balance < 0.9; heterozygous genotypes with an allele balance < 0.25 or > 0.75. Variant sites were then excluded if they met any of the following criteria: labelled as ‘mono-allelic’^64^; > 6 alternative alleles (including indels and SNVs); no alternative alleles after applying genotype QC; within the X or Y chromosome; mean genotype quality across all samples ≤ 40; call rate ≤ 0.9; or overlapped a low-complexity region. In total, 19,724,617 variants remained following variant QC.

Sample level QC was undertaken to ensure only samples with high-quality sequencing data remained in the dataset. The number of samples excluded and retained following each QC step are presented in Supplementary Table 16. Participants were excluded if they met any of the following criteria: sequencing call rate < 0.8 (n = 234, Supplementary Figure 5); discordant self-reported sex, imputed sex, and y chromosome depth (n = 210, Supplementary Figure 6, see Supplementary Materials 1.5 for detail); diagnoses of schizophrenia, autism spectrum disorder, or intellectual disability from primary care data, hospital inpatient data, death register records, or self-report (n = 1,526); or insufficient data for inferring ancestries (lacking or low quality array data, n = 1,293). No sample was excluded for low mean sequencing depth or mean genotype quality, most likely because this dataset had already received central QC prior to public release. To identify related individuals, we used estimates of pair-wise kinship co-efficients from array data. The ‘*ukb_gen_samples_to_remove’* command from the R package ukbtools^51^ was used to remove 69,274 samples ensuring that no pairs of retained participants were third degree relations or closer, defined as a kinship co-efficient > 0.0442^65,66^. In total, 72,537 individuals failed sample QC, retaining 396,848 individuals for analysis.

Analyses for inferring ancestry are based on the procedure described in^67^ and updated in^68^, and were conducted using UKBB array data. A description of this process is provided in detail in Supplementary Materials 1.1. In short, array data was used alongside biogeographical categories based on a standardised system^69^ and a 1KGP Phase 3 reference^30^ to infer Ancestry Informative Markers. PC-air was used to compute principal components in the 1KGP reference population, and project UKBB samples into the reference PC co-ordinates. The Tracy-Widom test^70^ selected eigenvectors for ancestry inference and a model based on Fisher’s Linear Discriminant Analysis^71^ was trained on known ancestries of the reference samples, and then applied to the input dataset to predict genetic ancestry probabilities. Genetic ancestry labels were then assigned based on these probabilities. We note that these labels merely reflect genetic similarity to the 1KGP reference panel^30^ and do not represent discrete or biologically distinct categories.

#### Variant annotation

RCVs, comprising single-nucleotide variants and insertions and deletions < 50 base-pairs in length, were annotated in Hail using Ensembl’s VEP^49^. PTVs were defined as splice acceptor, splice donor, stop-gain or frameshift variants that were annotated as high confidence for causing loss of protein function by LoFTEE^31,32^. Deleterious missense variants were defined as missense variants with a REVEL^33^ score > 0.75. This REVEL score threshold was chosen based on specificity for predicting deleterious variants in the Human Gene Mutation Database^33^. We also examined rare synonymous variants as a negative control.

#### Schizophrenia-associated gene sets

All gene sets are described in detail in Supplementary Table 5. Genes under selective constraint for PTVs, known as LoFi genes, are enriched for damaging RCVs, rare CNVs, and common alleles in schizophrenia^13–15^. We therefore analysed RCVs in 3,162 autosomal LoFi genes (defined as genes with a GnomAD pLI score^34^ ≥ 0.9) and in the remaining LoF-tolerant genes (those with a GnomAD pLI score < 0.9). Many schizophrenia-associated gene sets are enriched for genes expressed in the brain. We therefore investigated RCVs in brain expressed genes, defined as genes with an average of > 5 fragments per kilobase of exon model per million mapped reads in the brain in an RNA sequencing study^72^.

To provide robust evidence that the biology of schizophrenia overlaps with that for cognitive function, we also tested the following sets of protein-coding, autosomal genes implicated in schizophrenia in the largest GWAS^16^, CNV^17^ and RCV^19^ studies of the disorder to date.

##### RCV enriched genes

Genes enriched for RCVs in schizophrenia were taken from the SCHEMA study^19^. We analysed 29 autosomal genes enriched for RCVs in schizophrenia at a FDR of < 5%.

##### Common allele loci

Genes implicated in schizophrenia by common alleles were taken from the largest schizophrenia GWAS^16^. This study identified 282 autosomal loci associated with schizophrenia and then prioritised credible causal protein-coding genes for some of these loci using fine-mapping and SMR supplemented by chromatin conformation analysis. We analysed three schizophrenia common allele loci gene sets: **1)** all 1,715 genes that overlapped the 282 implicated loci; **2)** 186 genes closest to the index SNP for each associated locus (loci where the nearest gene to the index SNP was non-coding were not included in this analysis); **3**) 101 credible causal protein-coding genes that were prioritised in the original paper^16^.

##### Schizophrenia CNV loci

A set of 63 CNVs for which there is at least some evidence for association with developmental disorders have been shown to be collectively enriched in people with schizophrenia^17^. We focussed our CNV analysis on 886 genes in regions defined by these 63 CNVs, that we refer to as the schizophrenia-enriched CNV set. 13 of these 63 CNVs have individually been associated with schizophrenia with robust statistical significance^17,18^, which we refer to as the schizophrenia-associated CNV set (n = 178 genes). To refine which genes within the schizophrenia-enriched CNV set may be contributing to *g*, additional exploratory analyses were performed by testing RCVs only in the schizophrenia-associated CNV loci, and also by testing LoFi and LoF-tolerant genes in the schizophrenia-enriched and schizophrenia-associated CNV sets separately (see Supplementary Table 5 for further details).

### Array data

UKBB participants were genotyped using the Affymetrix Axiom UKBB array (∼450,000 individuals) and the UK BiLEVE array (∼50,000 individuals). Schizophrenia PRS were derived via a clumping and thresholding approach in PRSice-2^48^ using de-duplicated summary statistics from the largest GWAS of schizophrenia to date^16^, as previously described^73,74^. Schizophrenia-enriched CNVs were called as previously described^21^.

### Phenotypic data

Phenotypic data were collected from UKBB participants at multiple timepoints, in person and online. All participants attended an initial assessment centre where baseline data were collected by: touchscreen questionnaires, including cognitive function measures which took around 5 minutes; a face-to-face interview; and blood sample collection. Participants were later invited to attend additional visits and to complete online questionnaires, including online cognitive test batteries. Where cognitive tests were completed at multiple timepoints, we selected the instance with the greatest sample size for use in our study. Supplementary Table 2 provides an overview of the cognitive tests examined in this study and the number of participants who completed them. For all tests, scores were converted to a normal distribution if not already normally distributed and standardised through conversion to z-scores. Outlying scores (outside of +\− 4 standard deviations of the mean) were excluded for each cognitive test.

#### Generalised cognition (g)

Cognitive assessments were brief and unsupervised. Individual cognitive tests are known to vary in their reliability and stability^36,75^, but performance between different cognitive tests is correlated^75,76^ (Supplementary Table 3). The latter property enables *g* to be derived from a PC analysis of multiple cognitive tests^36^. We derived *g* as the first PC from a PC analysis of the following: **1)** Numeric memory (online test battery 1), **2)** Reaction time, **3)** Pairs matching, **4)** Trail making test B (online test battery 1). A detailed description of how *g* was derived, and the rationale for the inclusion/exclusion of individual cognitive tests in *g*, are provided in Supplementary Materials 1.2. In total, we had sufficient data to calculate *g* for 76,783 participants.

### Statistical analysis

Linear regression was used to test for association between *g*, the dependent variable, and RCV burden in different schizophrenia-associated gene sets and individual genes. We included as covariates burden of synonymous variants (apart from when investigating synonymous variant burden), sequencing batch and assessment centre. Given cognitive ability is non-linearly associated with age, and that this can differ by sex, we also included covariates for sex and standardised age, standardised age^2^, sex*standardised age and sex* standardised age^2^.

Additionally, we included covariates to control for population stratification, including ancestry probabilities (excluding 1KGP-EUR-like probability, as if all ancestry probabilities were included these would be co-linear), and relevant within ancestry PCs 1-10 (e.g. for analysis of the 1KGP-EUR-like population, the first 10 PCs derived from just this population were included as covariates). For the analysis of the participants who were assigned the ‘Admixed’ 1KGP-like genetic ancestry label (see Supplementary Materials 1.1), we covaried for PCs derived in all participants rather than for within ancestry PCs. All reported regression p-values are two-sided. We explored whether associations of RCV burden and *g* were explained by variation in specific cognitive tests by including individual cognitive test scores as covariates in the analyses of LoFi genes.

A multivariable linear regression model was used to test whether damaging RCVs, schizophrenia PRS, and schizophrenia-enriched CNVs had independent effects on cognition. Here, we included all covariates used in the analysis of rare coding variation alone, as well as an additional covariate for genotyping array. This analysis was run in 75,111 individuals of 1KGP-EUR-like ancestries in whom *g* could be derived and who had high-quality data for all classes of genetic variation investigated.

To compare associations of RCV burden and *g* between gene sets, we performed z-tests using the equation 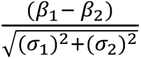 where: *β*_1_ is the beta and *σ*_1_ is the standard error of association from the RCV burden in gene set 1; and *β*_2_ is the beta and *σ*_2_ is the standard error of association from the RCV burden in gene set 2. Only independent gene sets were compared using z-tests.

## Acknowledgments

We thank George Kirov for contributions to CNV calling, Antonio Pardiñas for developing the pipeline from which we worked to infer genetic ancestries, and Sophie Legge for performing quality control of the array data and deriving schizophrenia polygenic risk scores.

This work was supported by a UKRI MRC Programme Grant (MR/Y004094/1) to JTRW, ER, PH, MCOD and MJO, a Wellcome Trust Integrative Neuroscience PhD Studentship to EF (108891/B/15/Z /WT), a UKRI Future Leaders Fellowship Grant to ER (MR/T018712/1 and MR/Y033922/1).

This research has been conducted using the UK Biobank Resource under Application Number 13310. This work uses data provided by patients and collected by the NHS as part of their care and support: Copyright © (2023), NHS England; Re-used with the permission of the NHS England and UK Biobank; All rights reserved.

This research used data assets made available by National Safe Haven as part of the Data and Connectivity National Core Study, led by Health Data Research UK in partnership with the Office for National Statistics and funded by UK Research and Innovation (research which commenced between 1st October 2020 – 31st March 2021 (MC_PC_20029); 1st April 2021 - 30th September 2022 (MC_PC_20058).

## Author contributions

E.F., E.R., and J.T.R.W. conceived and designed the research. E.F. and E.R. analysed the data. E.F., E.R., J.T.R.W., P.H., M.C.O.D. and M.J.O. contributed to the interpretation of the results. E.F., E.R., and J.T.R.W. wrote the manuscript, which was read, edited and approved by all authors.

## Resource Availability

### Lead contact

Requests for further information and resources should be directed to and will be fulfilled by the lead contact, Dr. Elliott Rees.

### Data availability

This study used data from the UK Biobank, which is available for health-related research upon registration and application through the UK Biobank Access Management System (https://www.ukbiobank.ac.uk/enable-your-research/register). The code required to reproduce our analyses is publicly available (https://github.com/eilidhfenner/UKBB_RAP_WES).

## Supplemental information

**Supplementary_Materials.docx :** Supplementary Materials and Supplementary Figures 1-6.

**Supplementary_Tables.xlsx :** Supplementary Tables 1-16.

## References

1. Saha, S., Chant, D., Welham, J. & McGrath, J. A Systematic Review of the Prevalence of Schizophrenia. PLOS Med. 2, e141 (2005).

2. Perälä, J. et al. Lifetime prevalence of psychotic and bipolar I disorders in a general population. Arch. Gen. Psychiatry 64, 19–28 (2007).

3. Sørensen, H. J., Mortensen, E. L., Parnas, J. & Mednick, S. A. Premorbid Neurocognitive Functioning in Schizophrenia Spectrum Disorder. Schizophr. Bull. 32, 578–583 (2006).

4. Reichenberg, A. et al. Static and dynamic cognitive deficits in childhood preceding adult schizophrenia: a 30-year study. Am. J. Psychiatry 167, 160–169 (2010).

5. Fusar-Poli, P. et al. Cognitive Functioning in Prodromal Psychosis: A Meta-analysis. Arch. Gen. Psychiatry 69, 562–571 (2012).

6. Bowie, C. R., Reichenberg, A., Patterson, T. L., Heaton, R. K. & Harvey, P. D. Determinants of Real-World Functional Performance in Schizophrenia Subjects: Correlations With Cognition, Functional Capacity, and Symptoms. Am. J. Psychiatry 163, 418–425 (2006).

7. Green, M. F., Horan, W. P. & Lee, J. Nonsocial and social cognition in schizophrenia: current evidence and future directions. World Psychiatry 18, 146–161 (2019).

8. Fioravanti, M., Carlone, O., Vitale, B., Cinti, M. E. & Clare, L. A Meta-Analysis of Cognitive Deficits in Adults with a Diagnosis of Schizophrenia. Neuropsychol. Rev. 15, 73–95 (2005).

9. Keefe, R. S. E. et al. The Brief Assessment of Cognition in Schizophrenia: reliability, sensitivity, and comparison with a standard neurocognitive battery. Schizophr. Res. 68, 283–297 (2004).

10. Pietrzak, R. H. et al. A comparison of the CogState Schizophrenia Battery and the Measurement and Treatment Research to Improve Cognition in Schizophrenia (MATRICS) Battery in assessing cognitive impairment in chronic schizophrenia. J. Clin. Exp. Neuropsychol. 31, 848–859 (2009).

11. Green, M. F., Kern, R. S., Braff, D. L. & Mintz, J. Neurocognitive Deficits and Functional Outcome in Schizophrenia: Are We Measuring the “Right Stuff”? Schizophr. Bull. 26, 119–136 (2000).

12. Dickinson, D., Iannone, V. N., Wilk, C. M. & Gold, J. M. General and specific cognitive deficits in schizophrenia. Biol. Psychiatry 55, 826–833 (2004).

13. Singh, T. et al. The contribution of rare variants to risk of schizophrenia in individuals with and without intellectual disability. Nat. Genet. 49, 1167–1173 (2017).

14. Rees, E. et al. Schizophrenia, autism spectrum disorders and developmental disorders share specific disruptive coding mutations. Nat. Commun. 12, 5353 (2021).

15. Pardiñas, A. F. et al. Common schizophrenia alleles are enriched in mutation-intolerant genes and in regions under strong background selection. Nat. Genet. 50, 381–389 (2018).

16. Trubetskoy, V. et al. Mapping genomic loci implicates genes and synaptic biology in schizophrenia. Nature 604, 502–508 (2022).

17. Rees, E. et al. Analysis of Intellectual Disability Copy Number Variants for Association With Schizophrenia. JAMA Psychiatry 73, 963–969 (2016).

18. Marshall, C. R. et al. Contribution of copy number variants to schizophrenia from a genome-wide study of 41,321 subjects. Nat. Genet. 49, 27–35 (2017).

19. Singh, T. et al. Rare coding variants in ten genes confer substantial risk for schizophrenia. Nature 1–9 (2022) doi:10.1038/s41586-022-04556-w.

20. Liu, D. et al. Schizophrenia risk conferred by rare protein-truncating variants is conserved across diverse human populations. Nat. Genet. 55, 369–376 (2023).

21. Kendall, K. M. et al. Cognitive Performance Among Carriers of Pathogenic Copy Number Variants: Analysis of 152,000 UK Biobank Subjects. Biol. Psychiatry 82, 103–110 (2017).

22. Stefansson, H. et al. CNVs conferring risk of autism or schizophrenia affect cognition in controls. Nature 505, 361–366 (2014).

23. Chen, C.-Y. et al. The impact of rare protein coding genetic variation on adult cognitive function. Nat. Genet. 1–12 (2023) doi:10.1038/s41588-023-01398-8.

24. Kingdom, R., Beaumont, R. N., Wood, A. R., Weedon, M. N. & Wright, C. F. Genetic modifiers of rare variants in monogenic developmental disorder loci. Nat. Genet. 56, 861–868 (2024).

25. Lencz, T. et al. Molecular genetic evidence for overlap between general cognitive ability and risk for schizophrenia: a report from the Cognitive Genomics consorTium (COGENT). Mol. Psychiatry 19, 168–174 (2014).

26. Mallet, J., Le Strat, Y., Dubertret, C. & Gorwood, P. Polygenic Risk Scores Shed Light on the Relationship between Schizophrenia and Cognitive Functioning: Review and Meta-Analysis. J. Clin. Med. 9, 341 (2020).

27. Gardner, E. J. et al. Reduced reproductive success is associated with selective constraint on human genes. Nature 603, 858–863 (2022).

28. Nuechterlein, K. H. et al. Identification of separable cognitive factors in schizophrenia. Schizophr. Res. 72, 29–39 (2004).

29. Schaefer, J., Giangrande, E., Weinberger, D. R. & Dickinson, D. The global cognitive impairment in schizophrenia: Consistent over decades and around the world. Schizophr. Res. 150, 42–50 (2013).

30. Auton, A. et al. A global reference for human genetic variation. Nature 526, 68–74 (2015).

31. Karczewski, K. J. et al. The mutational constraint spectrum quantified from variation in 141,456 humans. Nature 581, 434–443 (2020).

32. Karczewski, K. konradjk/loftee. (2020).

33. Ioannidis, N. M. et al. REVEL: An Ensemble Method for Predicting the Pathogenicity of Rare Missense Variants. Am. J. Hum. Genet. 99, 877–885 (2016).

34. Lek, M. et al. Analysis of protein-coding genetic variation in 60,706 humans. Nature 536, 285–291 (2016).

35. McCutcheon, R. A., Keefe, R. S. E. & McGuire, P. K. Cognitive impairment in schizophrenia: aetiology, pathophysiology, and treatment. Mol. Psychiatry 1–17 (2023) doi:10.1038/s41380-023-01949-9.

36. Fawns-Ritchie, C. & Deary, I. J. Reliability and validity of the UK Biobank cognitive tests. PLoS ONE 15, (2020).

37. Johnson, W., Nijenhuis, J. te & Bouchard, T. J. Still just 1 g: Consistent results from five test batteries. Intelligence 36, 81–95 (2008).

38. Johnson, W., Bouchard, T. J., Krueger, R. F., McGue, M. & Gottesman, I. I. Just one g: consistent results from three test batteries. Intelligence 32, 95–107 (2004).

39. Kennedy, M. B. Synaptic Signaling in Learning and Memory. Cold Spring Harb. Perspect. Biol. 8, a016824 (2016).

40. MacLeod, A. K. et al. Genetic Copy Number Variation and General Cognitive Ability. PLOS ONE 7, e37385 (2012).

41. Smeland, O. B., Frei, O., Dale, A. M. & Andreassen, O. A. The polygenic architecture of schizophrenia — rethinking pathogenesis and nosology. Nat. Rev. Neurol. 16, 366–379 (2020).

42. Rolland, T. et al. Phenotypic effects of genetic variants associated with autism. Nat. Med. 1–10 (2023) doi:10.1038/s41591-023-02408-2.

43. Fry, A. et al. Comparison of Sociodemographic and Health-Related Characteristics of UK Biobank Participants With Those of the General Population. Am. J. Epidemiol. 186, 1026–1034 (2017).

44. Schoeler, T. et al. Participation bias in the UK Biobank distorts genetic associations and downstream analyses. *Nat*. Hum. Behav. 7, 1216–1227 (2023).

45. Hail Team. Hail. https://github.com/hail-is/hail.

46. Chang, C. C. et al. Second-generation PLINK: rising to the challenge of larger and richer datasets. GigaScience 4, s13742-015-0047–8 (2015).

47. Euesden, J., Lewis, C. M. & O’Reilly, P. F. PRSice: Polygenic Risk Score software. Bioinformatics 31, 1466–1468 (2015).

48. Choi, S. W. & O’Reilly, P. F. PRSice-2: Polygenic Risk Score software for biobank-scale data. GigaScience 8, giz082 (2019).

49. McLaren, W. et al. The Ensembl Variant Effect Predictor. Genome Biol. 17, 122 (2016).

50. Wickham, H. Ggplot2. (Springer International Publishing, Cham, 2016). doi:10.1007/978-3-319-24277-4.

51. Hanscombe, K. ukbtools: Manipulate and Explore UK Biobank Data. (2019).

52. Extension of ‘data.fram’. https://rdatatable.gitlab.io/data.table/.

53. A Grammar of Data Manipulation. https://dplyr.tidyverse.org/.

54. Convert Statistical Objects into Tidy Tibbles. https://broom.tidymodels.org/.

55. Van Hout, C. V. et al. Exome sequencing and characterization of 49,960 individuals in the UK Biobank. Nature 586, 749–756 (2020).

56. Poplin, R. et al. A universal SNP and small-indel variant caller using deep neural networks. Nat. Biotechnol. 36, 983–987 (2018).

57. Szustakowski, J. D. et al. Advancing human genetics research and drug discovery through exome sequencing of the UK Biobank. Nat. Genet. 53, 942–948 (2021).

58. Regier, A. A. et al. Functional equivalence of genome sequencing analysis pipelines enables harmonized variant calling across human genetics projects. Nat. Commun. 9, 4038 (2018).

59. Krasheninina, O. et al. Open-source mapping and variant calling for large-scale NGS data from original base-quality scores. 2020.12.15.356360 Preprint at 10.1101/2020.12.15.356360 (2020).

60. dnanexus/oqfe. https://hub.docker.com/r/dnanexus/oqfe.

61. GRCh38 - hg38 - Genome - Assembly - NCBI. https://www.ncbi.nlm.nih.gov/assembly/GCF_000001405.26/.

62. UKB Showcase: WES protocol. https://biobank.ndph.ox.ac.uk/showcase/refer.cgi?id=915.

63. UKB Showcase: Exome Sequences. https://biobank.ndph.ox.ac.uk/showcase/label.cgi?id=170.

64. Lin, M. F., et al. GLnexus: joint variant calling for large cohort sequencing. bioRxiv 343970 (2018) doi:10.1101/343970.

65. Relationship Inference in KING. https://www.kingrelatedness.com/manual.shtml.

66. Manichaikul, A. et al. Robust relationship inference in genome-wide association studies. Bioinformatics 26, 2867–2873 (2010).

67. Legge, S. E. et al. A genome-wide association study in individuals of African ancestry reveals the importance of the Duffy-null genotype in the assessment of clozapine-related neutropenia. Mol. Psychiatry 24, 328–337 (2019).

68. Smart, S. E. et al. SLC39A8.p.(Ala391Thr) is associated with poorer cognitive ability: a cross-sectional study of schizophrenia and the general UK population. 2024.09.18.24313865 Preprint at 10.1101/2024.09.18.24313865 (2024).

69. Huddart, R. et al. Standardized Biogeographic Grouping System for Annotating Populations in Pharmacogenetic Research. Clin. Pharmacol. Ther. 105, 1256–1262 (2019).

70. Patterson, N., Price, A. L. & Reich, D. Population Structure and Eigenanalysis. PLOS Genet. 2, e190 (2006).

71. Yang, N. et al. Examination of ancestry and ethnic affiliation using highly informative diallelic DNA markers: application to diverse and admixed populations and implications for clinical epidemiology and forensic medicine. Hum. Genet. 118, 382–392 (2005).

72. Fagerberg, L. et al. Analysis of the human tissue-specific expression by genome-wide integration of transcriptomics and antibody-based proteomics. Mol. Cell. Proteomics MCP 13, 397–406 (2014).

73. Legge, S. E. et al. Association of Genetic Liability to Psychotic Experiences With Neuropsychotic Disorders and Traits. JAMA Psychiatry 76, 1256–1265 (2019).

74. Legge, S. E. et al. Genetic and Phenotypic Features of Schizophrenia in the UK Biobank. JAMA Psychiatry 81, 681–690 (2024).

75. Lyall, D. M. et al. Cognitive Test Scores in UK Biobank: Data Reduction in 480,416 Participants and Longitudinal Stability in 20,346 Participants. PLoS ONE 11, e0154222 (2016).

76. Spearman, C. ‘General Intelligence’ Objectively Determined and Measured. 73 (Appleton-Century-Crofts, East Norwalk, CT, US, 1961). doi:10.1037/11491-006.

