## Supplementary Materials and Figures for "Rare coding variants in schizophrenia-associated genes affect generalised cognition in the UK Biobank"

#### 1.1 Genetic ancestry inference

Analyses for inferring genetic ancestry are based on the procedure described in[1] and updated in[2], and were conducted using UKBB array data. This data was first quality controlled using PLINK as in[3]. Single-nucleotide variants (SNVs) were excluded if they met any of the following: minor allele frequency (MAF) < 0.01; Hardy-Weinberg equilibrium $P$ value < $1.00 \times 10^{-6}$ (using the midp and keep-fewhet options for multi-population datasets); imputation quality information score < 0.9; SNV call rate < 0.95; within the MHC region; or within long-range high LD regions[4]. Individuals with SNV missingness > 0.05 were excluded. The post-QC dataset was formed of 487,409 samples and included 7,074,483 variants. This data was used alongside biogeographical categories based on a standardised system[5] and a 1000 Genomes Phase 3 (1KGP) reference[6] to infer 65,338 Ancestry Informative Markers (AIMs). AIMs are variants which differ greatly in terms of allele frequencies across biogeographical genetic ancestries. PC-air was used to compute the first 100 principal components (PCs) in the reference (1KGP) population, and project the UKBB samples into the reference PC co-ordinates.

The Tracy-Widom test[7] selected the 34 first eigenvectors of the dataset for the ancestry inference procedure. A model based on Fisher's Linear Discriminant Analysis (LDA)[8] was trained on the known ancestries of the reference samples with principal components as predictors. The trained LDA classification model was then applied to the principal components of the input dataset to predict the most likely genetic ancestry category of each sample.

Genetic ancestry labels were assigned based on genetic ancestry probabilities, which reflect genetic similarity to the 1KGP reference panel used[6]. For any individual, if any genetic ancestry probability was > 0.8, we assigned the 1KGP-ancestry-like label to that individual. We note that these labels merely reflect genetic similarity to the reference population and do not represent discrete or biologically distinct categories. Where no genetic ancestry probabilities were > 0.8, we assigned an 'Admixed' label to the individual. Sample sizes of each 1KGP-like genetic ancestry are presented in Supplementary Table 1.

For each genetic ancestry label, we derived 10 ancestry-specific PCs using UKBB post-QC array data for all unrelated participants with that label. Further quality control processes were applied: variants were pruned twice in PLINK to ensure limited linkage disequilibrium between SNPs ($R^2 > 0.2$); and rare ($MAF \leq 0.05$) variants were excluded. PC analysis was then run on remaining variants in Hail to output 10 ancestry-specific PCs for use as covariates in ancestry-specific analyses.

#### 1.2 Deriving generalised cognition (*g*)

We derived *g* from the first principal component (PC) from a PC analysis of the following cognitive tests: **1)** Numeric memory (online test battery 1), **2)** Reaction time (baseline assessment), **3)** Pairs matching (baseline assessment), **4)** Trail making test B (online test battery 1). *g* was only available for 76,783 participants who completed all four input tests. In selecting tests for use in the formation of *g*, we aimed to find a combination of four tests which each spanned different cognitive domains, whilst maximising sample size. Tests with low completion rates (<20%) were thus not considered for inclusion. We also aimed to select tests administered as close together in time as possible, and therefore did not consider tests completed in the second online cognitive test battery for inclusion in *g*, as this battery was completed 11 or more years after the initial assessment centre. Our estimates of *g* were standardised, and outliers above and below 4 standard deviations from the mean were removed.

#### 1.3 Additional measures of *g* in the UK Biobank

To investigate the impact of modifying some of the cognitive tests included in the formation of *g*, we compared four separate *g* scores that were formed using three of the four tests included in our original estimate of *g*, leaving one test out at a time. All *g* scores were coded so a positive score reflected better performance on the input tests. Each of these *g* scores was highly correlated with the original *g* (weakest correlation co-efficient = 0.88, Supplementary Table 4), suggesting our measure of *g* is consistent with other possible measures, and it is not driven by any individual cognitive test.

In some studies of *g* in the UKBB, fluid intelligence is included in the formation of *g*. To investigate how similar our measure of *g* is compared to other possible measures, we formed two additional *g* scores: **1)** adding fluid intelligence to the PCA used to form our original *g*; **2**) using the five cognitive tests (pairs matching, reaction time, prospective memory, fluid intelligence and numeric memory) used to form *g* in the UKBB in Fawns-Ritchie et. al[9] (which they termed '*g: UKB-5'*), which was shown to be highly correlated with an independent measure of *g* formed from a battery of standard, well-validated cognitive measures. Both of these measures of *g* were highly correlated with our original measure (respective correlation coefficients = 0.94 and 0.80, Supplementary Table 4), but they were only available in less than half the number of samples included in our original measure of *g*. Thus, to maximise our sample size, we did not use these alternative measures of *g* for our primary analyses.

#### 1.4 Ethics

All participants gave consent for their data to be used by UKBB projects and agreed to being followed up. Ethical approval was granted to UKBB by several committees as it spans England, Scotland, and Wales: The Northwest Multi-Centre Research Ethics Committee; The Patient Information Advisory Group, which has since been replaced by the National Information Governance Board for Health and Social Care; and The Community Health Index Advisory Group.

#### 1.5 Inferring genetic sex

To infer genetic sex, high quality, common variants on the X chromosome (original n variants = 425,743) were selected by filtering variants on call rate (> 0.97, n variants = 297,719) and MAF (> 1%, n variants = 677). These high quality, common variants were then used to calculate the inbreeding co-efficient (F-statistic) on the X chromosome using the '*impute_sex*' command in Hail. The Y chromosome (original n variants = 10,432) was filtered to high quality variants (non-PAR variants with a mean depth ≥ 3.5, n variants = 8,737). F-statistic and Y-depth were then used to infer genetic sex: participants with F-statistic ≤ 0.6 were classified as female (n = 215,336) and those with an F-statistic > 0.6 were classified as male (n = 184,541); participants with mean Y chromosome depth ≤ 3 were inferred to not have a Y chromosome (n = 215,363), and those with mean Y chromosome depth > 3 were inferred to have a Y chromosome (n = 184,514). These measures were used alongside reported sex (UKBB Field 31[10], sex either acquired from central registry or self-reported (215,344 (53.85%) female, and 184,533 (46.15%) male) to assign inferred sex. Concordance between reported sex, F-statistic inferred sex, and Y chromosome inference was required to identify samples as female (n = 215,235) or male (n = 184,432), and all other individuals were excluded from this analysis (n = 210, Supplementary Figure 6), as the data suggests these individuals have low quality sequencing data, chromosomal abnormalities, or are potential sample swaps.

#### Supplementary Figures

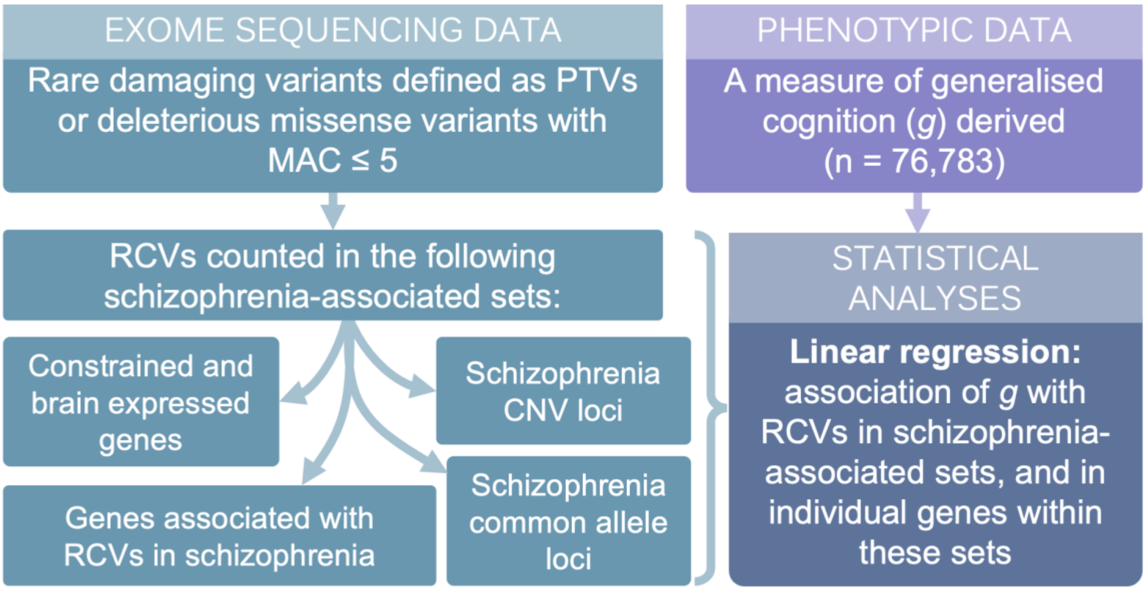

**Supplementary Figure 1**– Summary of analyses presented in this study. PTVs = protein truncating variants. MAC = minor allele count. RCVs = rare coding variants. CNV = copy number variant.

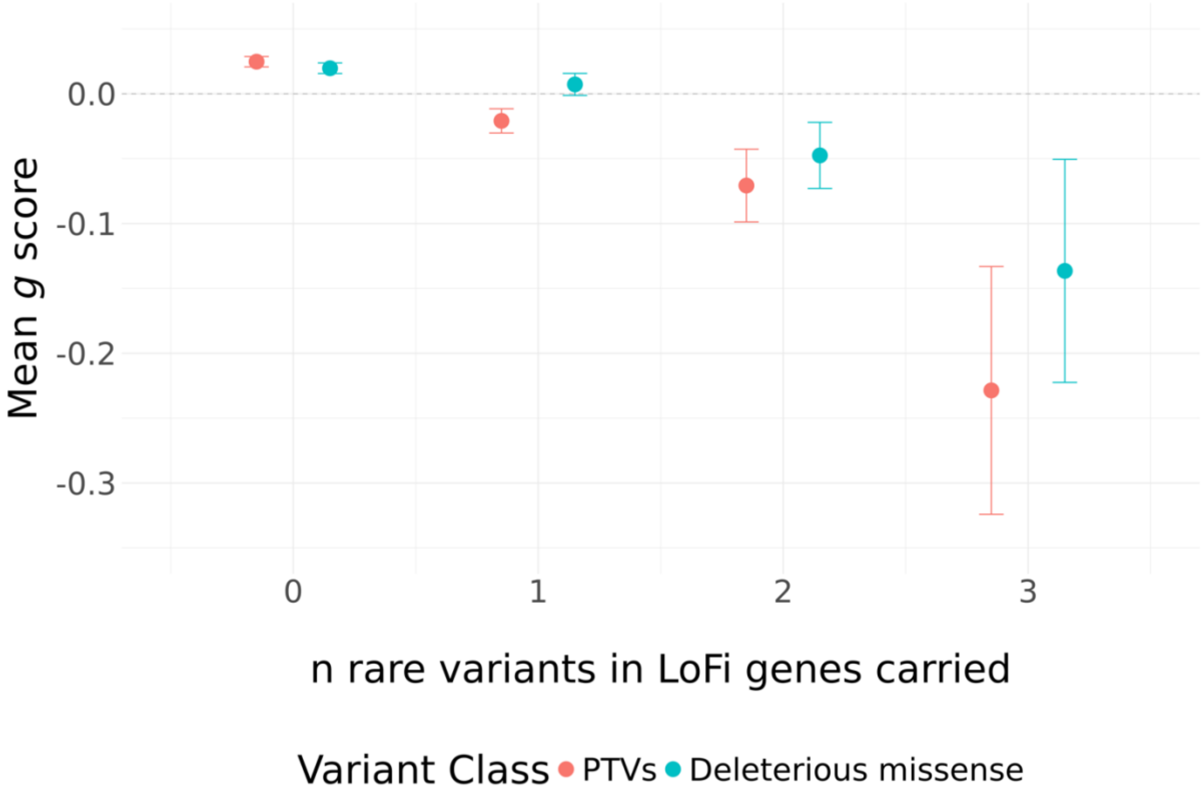

**Supplementary Figure 2** - The mean of *g* (a standardised measure) across individuals with different burden of rare protein truncating variants (PTVs) and rare deleterious missense variants in LoF-intolerant (LoFi) genes. < 10 participants carried ≥ 4 rare PTVs or deleterious missense variants in LoFi genes, and *g* could not be estimated accurately in this small a sample. These counts are therefore not plotted here. Error bars display 95% confidence intervals. PTV = protein-truncating variant; *g* = generalised cognition.

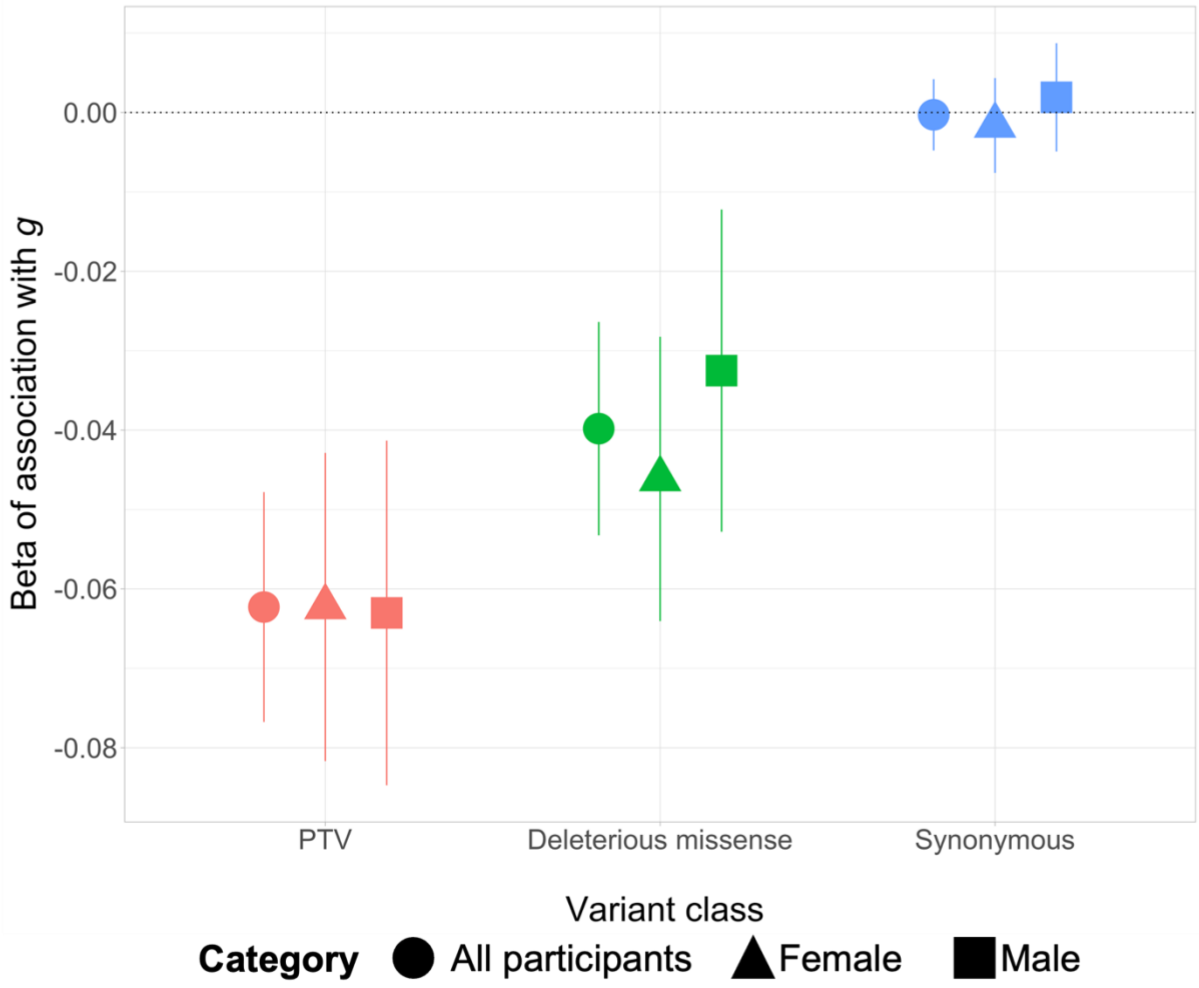

**Supplementary Figure 3 -** Association of *g* with different classes of rare variants in LoFi genes. Shapes represent the subset in which the analysis was run: all participants (circle, n = 75,188); females (triangle, n = 40,969); and males (squares, n = 34,219). Error bars display 95% confidence intervals. PTV = protein-truncating variant; *g* = generalised cognition.

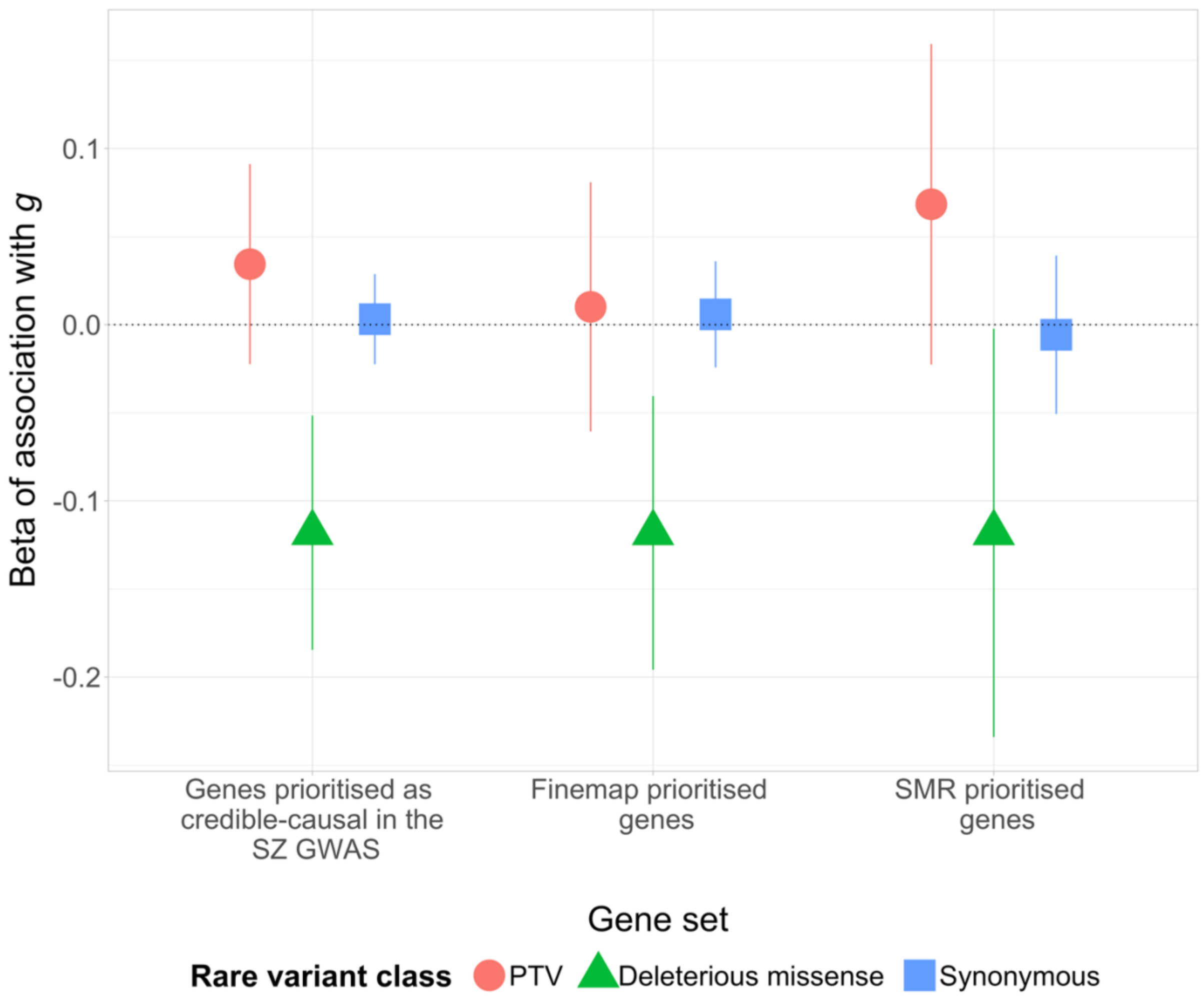

**Supplementary Figure 4 -** Association of *g* with different classes of rare variants in genes prioritised as credible causal by different methods in the schizophrenia (SZ) GWAS. Error bars display 95% confidence intervals. PTV = protein-truncating variant; *g* = generalised cognition.

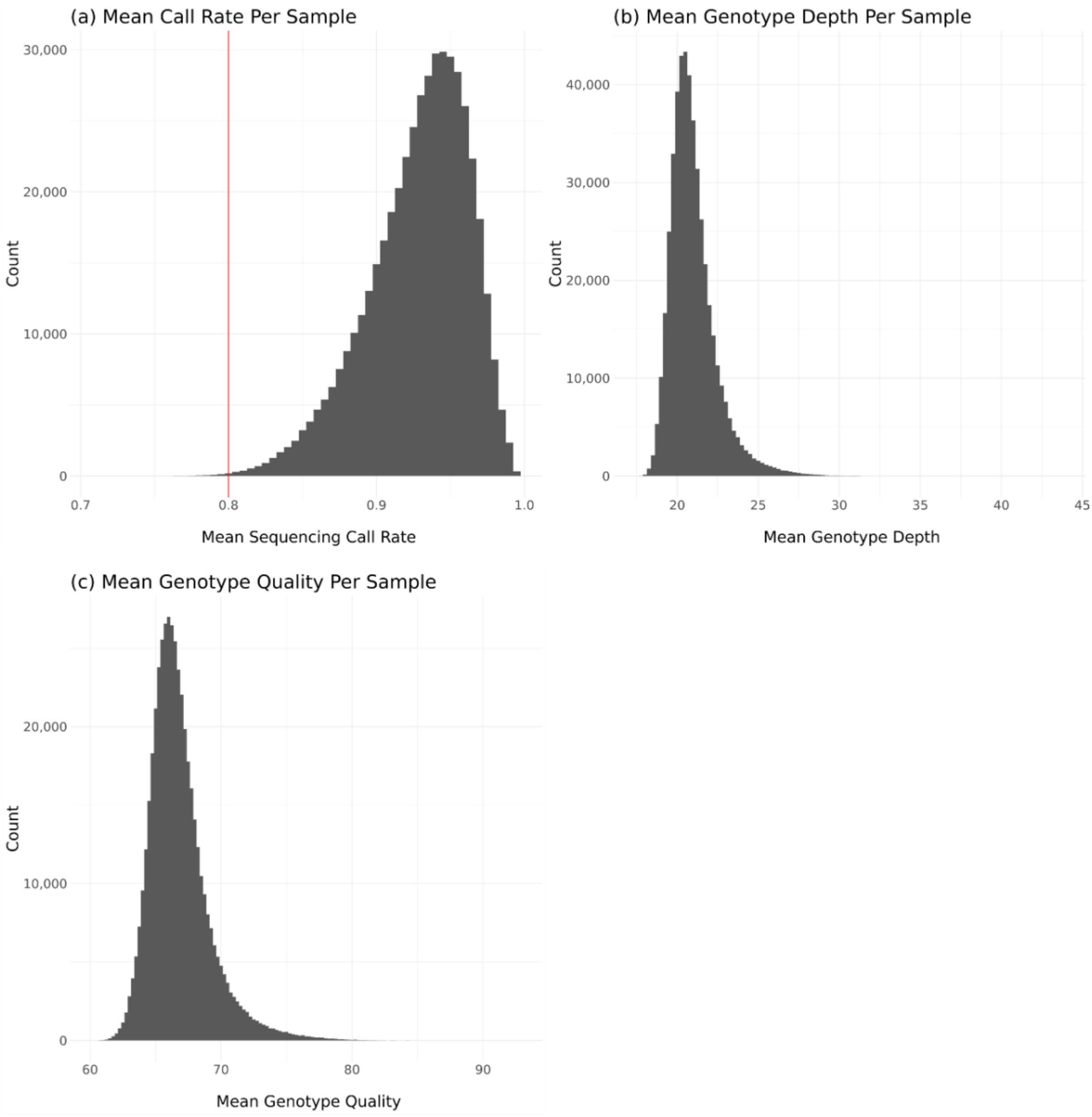

**Supplementary Figure 5 –** Plots of sample QC metrics following genotype-level quality control: a) mean call rate per sample, with a red line representing the cut-off applied at 0.8; b) mean genotype depth per sample; c) mean genotype quality per sample.

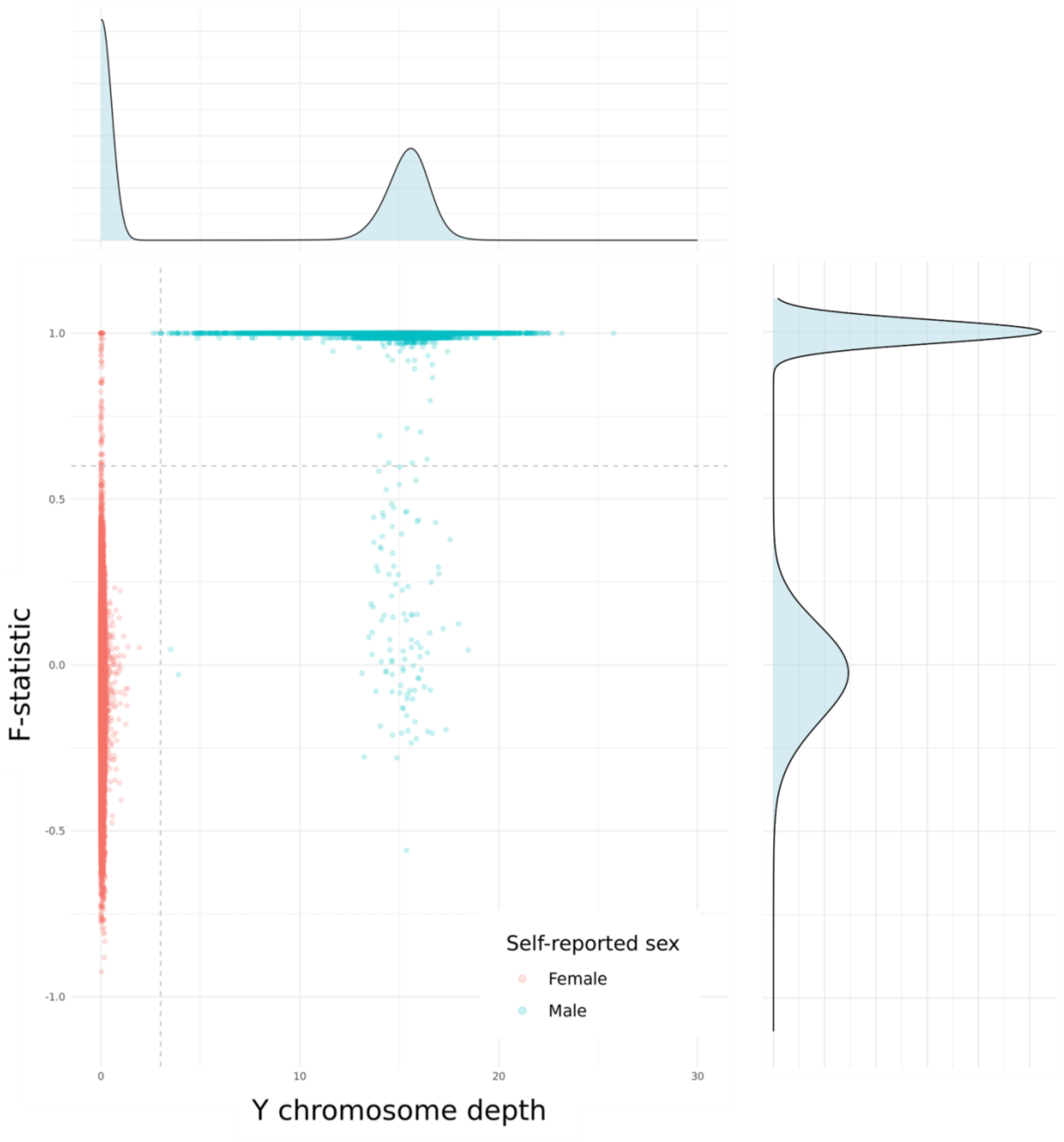

**Supplementary Figure 6** - F-statistic and mean Y chromosome variant mean depth for each participant. Participant who self-reported as male (blue dots) with F-statistics around 1 and non-zero Y chromosome depth were identified as male, and participants who self-reported as female (red dots) with low F-statistics and Y chromosome depth close to 0 were identified as female. Grey dotted lines reflect cut-offs used to define: F-statistic imputed sex (horizontal line at F-statistic = 0.6); and Y chromosome depth (vertical line at Y depth = 3).

### References

1. Legge, S. E. *et al.* A genome-wide association study in individuals of African ancestry reveals the importance of the Duffy-null genotype in the assessment of clozapine-related neutropenia. *Mol. Psychiatry* **24**, 328–337 (2019).

2. Smart, S. E. *et al.* SLC39A8.p.(Ala391Thr) is associated with poorer cognitive ability: a cross-sectional study of schizophrenia and the general UK population. 2024.09.18.24313865 Preprint at https://doi.org/10.1101/2024.09.18.24313865 (2024).

3. Legge, S. E. *et al.* Genetic and Phenotypic Features of Schizophrenia in the UK Biobank. *JAMA Psychiatry* **81**, 681–690 (2024).

4. Price, A. L. *et al.* Long-Range LD Can Confound Genome Scans in Admixed Populations. *Am. J. Hum. Genet.* **83**, 132–135 (2008).

5. Huddart, R. *et al.* Standardized Biogeographic Grouping System for Annotating Populations in Pharmacogenetic Research. *Clin. Pharmacol. Ther.* **105**, 1256–1262 (2019).

6. Auton, A. *et al.* A global reference for human genetic variation. *Nature* **526**, 68–74 (2015).

7. Patterson, N., Price, A. L. & Reich, D. Population Structure and Eigenanalysis. *PLOS Genet.* **2**, e190 (2006).

8. Yang, N. *et al.* Examination of ancestry and ethnic affiliation using highly informative diallelic DNA markers: application to diverse and admixed populations and implications for clinical epidemiology and forensic medicine. *Hum. Genet.* **118**, 382–392 (2005).

9. Fawns-Ritchie, C. & Deary, I. J. Reliability and validity of the UK Biobank cognitive tests. *PLoS ONE* **15**, (2020).

10.: Data-Field 31. https://biobank.ctsu.ox.ac.uk/crystal/field.cgi?id=31.
